# Syndemic Clustering of Noncommunicable Disease Risk Among Syrian Refugees and Jordanian Nationals: A Subnational Analysis of the WHO STEPS 2019 Survey

**DOI:** 10.1101/2025.08.01.25332574

**Authors:** Mannat Tiwana, Gursimran Singh Walia

## Abstract

Understanding the intersection of noncommunicable disease (NCD) risk and displacement in fragile settings is critical for equitable global health policy. This study employs a syndemic framework to investigate the clustering of behavioral risk factors—tobacco use, insufficient fruit intake, and physical inactivity—among Syrian refugees and Jordanian nationals, using data from the WHO STEPS 2019 survey. A composite risk score was constructed, and subnational analyses were conducted by sex and region. Among 2707 adults with complete data, 61.5% exhibited all three risk behaviors. Syrian refugees had a significantly higher prevalence of full-risk clustering (67.8%) than Jordanians (52.2%, p < 0.001), with the North and Center governorates displaying the greatest burden. Syrian women exhibited the highest syndemic risk. No statistically significant associations were found between the risk score and self-reported hypertension or diabetes. These findings underscore the need for gender-sensitive, subnationally targeted NCD strategies that address the structural drivers of health inequity in displacement-affected settings.

## Introduction

Noncommunicable diseases (NCDs) constitute the predominant cause of mortality globally, accounting for approximately 74% of all deaths, with the majority occurring in low- and middle-income countries [1]. Jordan is emblematic of this epidemiological transition, with NCDs contributing to 78% of national mortality [2]. Compounding this burden is the protracted displacement of Syrian refugees, who now represent a substantial proportion of Jordan’s population. The convergence of NCD risks with displacement-related vulnerability presents a critical public health challenge.

The concept of a syndemic offers a robust analytical lens to capture the compounded effect of co-occurring diseases and social inequities. Initially conceptualized by Singer et al. [5], syndemics underscore the interaction between social conditions and disease clustering. In contexts of forced migration, where systemic barriers to care, food insecurity, and gender inequity persist, the potential for syndemic amplification is considerable [6,7]. While the WHO STEPwise approach (STEPS) has been pivotal in capturing population-level NCD risk data, few studies have utilized these data to examine syndemic dynamics in refugee-hosting countries.

This study addresses that gap by exploring the extent and distribution of behavioral risk clustering across nationality, region, and sex in Jordan. Furthermore, it evaluates whether this clustering is associated with diagnosed hypertension or diabetes, thus informing both surveillance strategies and health system interventions.

## Methods

This analysis utilized data from the 2019 Jordan STEPS Survey, a nationally representative, cross-sectional dataset designed by the World Health Organization and the Jordanian Ministry of Health. The sample included 5713 adults aged 18–69 years, comprising both Jordanian nationals and Syrian refugees residing outside formal camp settings. Data collection followed the three STEPS modules: behavioral risk factors (Step 1), physical measurements (Step 2), and biochemical assessments (Step 3) [9].

For the present study, a composite syndemic risk score was derived from three behavioral variables: current tobacco use, insufficient fruit intake (defined as <3 days/week), and physical inactivity (<150 minutes of moderate activity per week). Each behavior was coded dichotomously and summed to create a score ranging from 0 to 3. Due to the low distribution of scores 0 and 1, analysis was limited to individuals scoring 2 or 3.

Descriptive statistics were employed to summarize the distribution of risk behaviors. Bivariate analyses, including Chi-square tests, assessed differences in risk score distribution across sex, nationality, and region. Further Chi-square tests examined associations between syndemic scores and self-reported hypertension and diabetes diagnoses. All analyses were conducted using SPSS.

Patients and members of the public were not involved in the design, conduct, reporting, or dissemination plans of this research. This study used secondary, de-identified data from a publicly available dataset collected through WHO and Ministry of Health protocols. The authors accessed the dataset on **June 13, 2025**, and had no access to any identifiable personal information at any stage of the analysis.

## Results

Of the 2707 respondents with complete risk behavior data, 1666 (61.5%) scored 3, indicating the presence of all three risk factors. The remaining 1041 (38.5%) scored 2. The mean syndemic risk score was 2.61. Syrian refugees were significantly more likely to exhibit full risk clustering (67.8%) than their Jordanian counterparts (52.2%) (p < 0.001). Geographic analysis showed that the Center and North regions bore the highest syndemic burden. Among Syrians residing in the North, 68.1% scored 3, compared to 58.5% of Jordanians in the same region. Similar trends were observed in the Center, where 67.6% of Syrians scored 3. These differences were statistically significant and highlight subnational disparities in behavioral NCD risk exposure.

Sex-disaggregated analysis revealed that Syrian women exhibited the highest level of syndemic clustering, with 69.2% scoring 3, compared to 61.0% of Jordanian women and 59.0% of Syrian men. These findings illustrate how gender intersects with nationality to shape NCD vulnerability.

Despite high levels of behavioral risk clustering, no significant associations were found between syndemic risk scores and self-reported hypertension (p = 0.696) or diabetes (p = 0.581). This disconnect may reflect underdiagnosis, poor access to screening, or delayed clinical manifestation of NCDs, particularly among refugees.

## Discussion

This study highlights critical syndemic dynamics among adults in Jordan, particularly among displaced Syrian populations and women. The high prevalence of behavioral clustering suggests urgent need for targeted health promotion interventions that are both geographically and demographically responsive.

The absence of significant associations between risk score and chronic disease diagnoses may not indicate a lack of disease burden but rather unmeasured systemic issues. These could include underdiagnosis due to limited access to care, diagnostic bias, or the cumulative nature of disease progression. These findings resonate with prior work documenting structural inequities in refugee health access in the EMR [6,13,14].

By applying a syndemic lens to national surveillance data, this study adds to a growing body of literature that challenges siloed approaches to NCD prevention [5,15,18]. It also supports calls for more nuanced NCD monitoring frameworks that recognize the intersectionality of gender, displacement, and structural deprivation [19,20].

## Conclusion

This study demonstrates that syndemic risk clustering is both widespread and unequally distributed across Jordan’s population, disproportionately affecting Syrian refugees and women in the North and Center regions. These findings advocate for equity-oriented health system reforms, including the integration of behavioral risk screening into routine refugee health services and the development of localized NCD prevention strategies.

## Acknowledgments

The author gratefully acknowledges the World Health Organization and the Jordanian Ministry of Health for providing open access to the WHO STEPS 2019 dataset.

## Competing Interests

The author declares no competing interests.

## Funding

This research did not receive any specific grant from funding agencies in the public, commercial, or not-for-profit sectors.

## Ethical Approval

The original STEPS survey protocol received ethical approval from the Jordanian Ministry of Health and the WHO Ethics Review Committee. The current study is a secondary analysis of this dataset and used de-identified, publicly available data, accessed on June 13, 2025. As such, no additional ethics approval was required.

## Data Availability

The dataset used in this study is publicly available from the World Health Organization STEPS website: https://www.who.int/data/data-collection-tools/steps

## Author Contributions

MT conceived the study, conducted the data analysis, interpreted the findings, and GSW wrote the manuscript.

## Notes

### Competing Interest Statement

The authors have declared no competing interest.

### Clinical Trial

NA

### Author Declarations

The original STEPS survey protocol received approval from the Jordanian Ministry of Health and the WHO Ethics Review Committee. The secondary analysis used de-identified, publicly available data and required no further ethical review.

